# The impact of anticholinergic burden on the development of mild behavioral impairment

**DOI:** 10.64898/2025.12.05.25341723

**Authors:** Clio E. Franklin, Paul B. Rosenberg, Constantine G. Lyketsos, Zahinoor Ismail, Jeannie-Marie Leoutsakos

**Affiliations:** Department of Psychiatry and Behavioral Sciences, Johns Hopkins University School of Medicine, 600 N Wolfe St, Baltimore, Maryland, 21287, USA; Department of Psychiatry, Mass General Brigham, 55 Fruit St, Boston, Massachusetts, 02114, USA; Departments of Psychiatry, Clinical Neurosciences, and Community Health Sciences, Hotchkiss Brain Institute, University of Calgary, 3330 Hospital Dr NW, Calgary, Alberta, T2N 4N1, Canada; NIHR Exeter Biomedical Research Centre, University of Exeter, St Luke’s Campus Heavitree Road, Exeter, EX1 2LU, UK

**Keywords:** Mild behavioral impairment, anticholinergic medication, neuropsychiatric symptoms, dementia, older adults

## Abstract

**Objective:** Mild Behavioral Impairment (MBI) is a syndrome of late-life-onset persistent neuropsychiatric symptoms. Anticholinergic medication is commonly prescribed in older adults. Both MBI and anticholinergic exposure are associated with increased dementia risk. We sought to understand the association of anticholinergic burden (ACB) with MBI.

**Design, Setting, participants:** We mapped ratings on the Neuropsychiatric Inventory Questionnaire to the MBI checklist (MBI-C) using an established algorithm to define MBI status in cognitively unimpaired individuals in the National Alzheimer’s Coordinating Center database. We then assessed the association between time-varying ACB ratings and risk of incident MBI.

**Results:** 4865 participants met inclusion criteria and were followed for a mean (SD) of 5.64 (3.92) years. ACB scores ranged from 0-11. 63.3% of participants had a score of 0, 27.7% had a score of 1-2, and 9% had a score of ≥3. Higher maximum total ACB score was associated with a higher likelihood of developing MBI (p=<0.001). When assessed as a time varying covariate, ACB score was associated with incident MBI (HR 1.12, 95% CI 1.05-1.19, p=<0.001). This association remained significant when adjusted for 10-year mortality risk.

**Conclusions:** MBI risk should be considered when prescribing anticholinergic medication in older adults.

## 1. Objective

Mild Behavioral Impairment (MBI) is a relatively new construct described by the International Society to Advance Alzheimer’s Research and Treatment (ISTAART) as ‘persistent neuropsychiatric symptoms (NPS) that emerge, *de novo*, in advance of or concurrent with Mild Cognitive Impairment (MCI)’.^1^ MBI symptoms can occur in ≥1 more of the following 5 domains: decreased motivation, emotional dysregulation, impulse dyscontrol, social inappropriateness, and abnormal perception or thought content.^1,2^ These domains have been quantified in the MBI Checklist (MBI-C), developed to more clearly characterize MBI symptoms and domains, as well as quantify MBI severity, thus better enabling clinicians and researchers to identify a group of patients in a “pre-dementia” state.^3^

MBI affects a significant proportion of older adults. A study of community dwelling, cognitively unimpaired (CU) older adults reported an MBI prevalence of 10%.^4^ A memory clinic study found an MBI prevalence of 37% in patients with subjective cognitive decline, and 54% in MCI.^5^ MBI can be a prodrome for cognitive decline, MCI or dementia.^4,6^ We reported that NPS and other behavioral symptoms precede cognitive manifestations of dementia in as many as 55% of participants.^6^ Others reported a 2.76-fold greater incidence rate of dementia in individuals with MBI when compared to those with no NPS.^7^ Individuals with MCI and concurrent MBI have lower reversion rates to CU and higher rates of progression to dementia.^8^

Alzheimer’s Dementia (AD) is predicted to affect 13.9 million Americans >65 years of age in 2060 with a 2025 cost burden of $781 billion in the USA. It is important to identify reversible risk factors for AD.^9,10^ Identifying variables linked to greater MBI risk may allow earlier recognition of individuals at higher risk for dementia as well as facilitate earlier interventions.

Medications with anticholinergic effects are recognized by the Beers Criteria as potentially inappropriate to use in older adults due to links with greater risks of side effects such as confusion, constipation, orthostasis, falls, and dry mouth.^11,12^ Anticholinergic medications are commonly prescribed to the elderly, with 9.1- 52.7% of individuals >65 years of age exposed to such medications.^13,14^ Anticholinergic prescribing is increasing: in >220,000 UK Biobank participants assessed between 1990-2015, anticholinergic burden, calculated by a ‘meta-scale’ combining 9 different anticholinergic ratings, increased 9-fold over that 15 year period.^15^

Long-term anticholinergic exposure is linked to greater dementia risk.^16,17^ A large, nested case-control study by Coupland et al. studied 284,343 General Practice patients in England ≥55 years of age. Compared to no anticholinergic drug prescriptions in the preceding 1-11 years, adjusted OR for dementia was 1.06 (95% CI 1.03-1.09) in the lowest anticholinergic exposure category versus 1.49 (95% CI 1.44-1.54) in the highest.^16^ This is supported by a case□control study of 395,397 Japanese individuals ≥65 years of age. Exposure to anticholinergic drugs led to significantly higher risk of developing dementia (adjusted OR 1.50, 95% CI 1.47-1.54).^17^

We are aware of no studies to date assessing the association between anticholinergic medication and development of MBI in CU subjects. Given that MBI can be a prodrome of dementia, establishing any impact of these medications on the onset of MBI could have a real-world impact on prescribing practices and guide future prospective studies.

## 2. Methods

### 2.1 Participants

Participants were volunteers with normal cognition recruited and then followed up approximately annually at Alzheimer’s Disease Research Centers (ADRCs), funded by the National Institute on Aging and located across the United States. The National Alzheimer’s Coordinating Center (NACC) maintains a database of ADRC participants’ neuropathologic and clinical data.^18^ To be included, individuals had to be age 65 and over, CU at baseline, and have a Neuropsychiatric Inventory Questionnaire (NPI-Q) score of 0 at their first and second ADRC visits.^19^ Those with neurodegenerative disorders (Huntington’s disease, Parkinson’s disease, amyotrophic lateral sclerosis, multiple system atrophy and progressive supranuclear palsy), developmental neuropsychiatric disorders (autism spectrum disorder, attention-deficit hyperactivity disorder), or Down Syndrome were excluded. Written informed consent was obtained from all participants at each ADRC and overseen by local IRBs.

Data collected by ADRCs accrue by NACC comprising a Uniform Data Set (UDS) which includes demographics, health history, medications, behavioral assessment, and neuropsychological battery.^20,21^ We examined data collected from ADRC visits between 6/2005 and 2/2020.

### 2.2 Measures

#### 2.2.1 Neuropsychiatric Inventory Questionnaire

The NPS assessment tool in the NACC database is the Neuropsychiatric Inventory questionnaire (NPI-Q). This is a widely used, validated, informant-based, self-report that evaluates the presence or absence of 12 NPS domains with a 0-3 rating of severity: delusions, hallucinations, agitation/aggression, depression/dysphoria, anxiety, elation/euphoria, apathy/indifference, disinhibition, irritability/lability, motor disturbance, night-time behaviors, and change in appetite/eating.^19^

#### 2.2.2 MBI diagnosis

To map the NPI-Q to the MBI-C we used the method of crosswalking described by Sheikh et al. shown in Table 1.^22^ A score of ≥1 in any NPI-Q domain resulted in a score in its corresponding MBI domain. To meet criteria for a diagnosis of MBI, a participant had to score ≥1 in any corresponding MBI domain, and their symptoms had to be persistent over a 6-month period. However, as the NPI-Q assesses symptoms over only the prior 1-month period, we used the TCV (two consecutive visits) MBI operational case definition. This considers MBI to be present if there is persistence of symptoms on the NPI-Q over two successive annual NACC visits.^23^

**Table 1.**
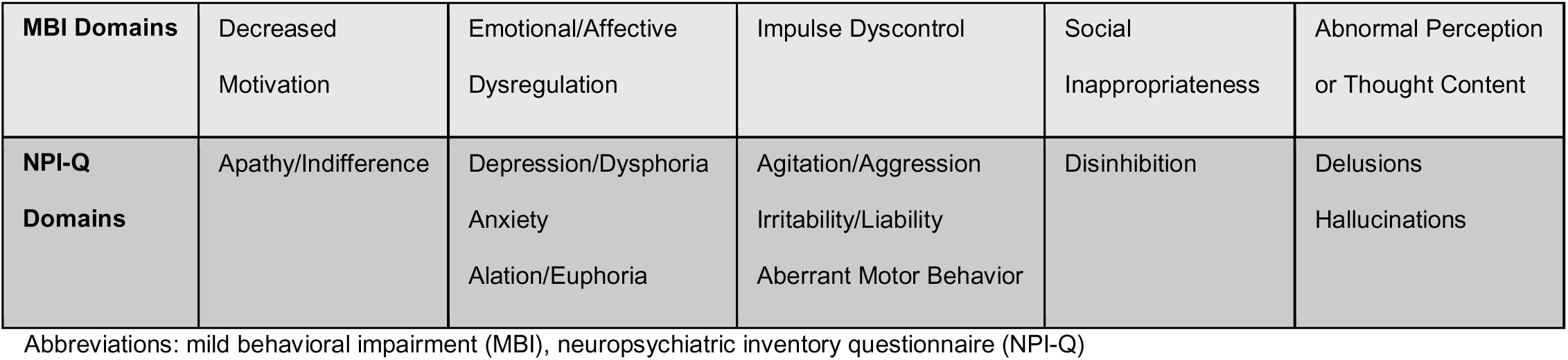
Mapping NPI-Q to MBI.

|  |  |  |  |  |  |
| --- | --- | --- | --- | --- | --- |
| **MBI Domains** | Decreased<br>Motivation | Emotional/Affective<br>Dysregulation | Impulse Dyscontrol | Social<br>Inappropriateness | Abnormal Perception<br>or Thought Content |
| **NPI-Q**<br>**Domains** | Apathy/Indifference | Depression/Dysphoria<br>Anxiety<br>Alation/Euphoria | Agitation/Aggression<br>Irritability/Liability<br>Aberrant Motor Behavior | Disinhibition | Delusions<br>Hallucinations |

Abbreviations: mild behavioral impairment (MBI), neuropsychiatric inventory questionnaire (NPI-Q)

#### 2.2.3 MCI and dementia diagnoses

Diagnoses were made by a consensus panel or an individual clinical assessment using all available UDS data.^24^ Modified Petersen criteria were used to diagnose MCI.^25,26^ NINCSD/ADRDA criteria were used to diagnose AD, the NINDS/AIREN criteria were used to diagnose vascular dementia, the Dementia with Lewy bodies consortium criteria were used to diagnose DLB, and consensus criteria as described in Neary et al. was used to diagnose frontotemporal dementia.^27-30^

#### 2.2.4 Anticholinergic Cognitive Burden scale

Individual anticholinergic medication exposure information was obtained from the NACC database, where each participant’s current medication list, including both prescribed and over the counter medication, is documented at their initial and each follow-up visit. To quantify anticholinergic exposure, we used the updated Anticholinergic Burden (ACB) scale.^31^ The ACB scale was developed in 2008 by researchers at the Indiana University Center for Aging and Regenstrief Institute for use as a tool to optimize geriatric pharmacotherapy by reducing anticholinergic burden. It was updated in 2012 with additional medications reviewed and added.^32^ Though multiple similar scales exist, the ACB was chosen as it lends itself to use in datasets such as the NACC, and has been widely used in other peer reviewed studies looking at anticholinergic burden.^33,34^

The ACB scale categorizes medications with a score of 1 (‘evidence from in vitro data that chemical entity has antagonist activity at muscarinic receptor’), 2 (‘evidence from literature, prescriber’s information, or expert opinion of clinical anticholinergic effect’), or 3 (‘evidence from literature, expert opinion, or prescriber’s information that medication may cause delirium’).^32^ If an individual was taking no anticholinergic medication, they received a score of 0. For a full list of medications and their ACB scores see supplemental table 1.

#### 2.2.6 Charlson Comorbidity Index

Charlson Comorbidity Index (CCI) calculates 10 year mortality risk by allocating a score to age and the following comorbidities: history of myocardial infarction, peripheral vascular disease, congestive heart failure, cerebrovascular disease, hemiplegia, chronic pulmonary disease, dementia, connective tissue disease, liver disease, peptic ulcer disease, diabetes, renal disease, tumor with or without metastases, leukemia, lymphoma, and AIDS.^35^ We included CCI in our multivariate model as a marker of participant morbidity.

### 2.3 Statistical methods

We categorized cognitive status as ‘CU’, ‘impaired but not MCI’, ‘MCI’, or ‘dementia’. Participants who developed MCI were not censored as MCI can be concurrently diagnosed with MBI.^1^ As described by Wise et al., a diagnosis of MCI or dementia was considered “sticky”, i.e., once a participant was diagnosed with dementia they were considered to have that diagnosis at subsequent visits and, once a participant was diagnosed with MCI they were considered to have that diagnosis at subsequent visits until they were diagnosed with dementia.^6^ Participants who developed dementia were censored at the time of their diagnosis.

To generate a Kaplan-Meier curve, each participant’s maximum ACB score across all visits was used.

In the primary analysis, ACB score was a time-varying covariate with participant scores calculated by adding together the ACB score of each medication they were taking at each ADRC visit during the study period. We fit a series of Cox proportional hazard models for hazard of MBI, with ACB score, non-aspirin NSAIDs, opioids, benzodiazepines, selective serotonin reuptake inhibitors (SSRIs), non-SSRI antidepressants, antipsychotics, aspirin, diphenhydramine, and oxybutynin as time-varying predictors. Psychoactive medication classes were chosen to assess for protopathic bias. In addition, there is some evidence to suggest that SSRIs may be neuroprotective.^36^ Benadryl and oxybutynin were chosen as they are commonly used medications with strong anticholinergic activity. Aspirin was chosen as a proxy marker of cardiovascular risk. We completed unadjusted models as well as models adjusting for age at baseline, sex, race, years of education, and CCI. We tested the assumption of proportional hazards via Schonfeld residuals.^37^

## 3. Results

4865 participants met inclusion criteria. Table 2 shows their baseline characteristics. Most participants were white females, who were well educated. Participants were followed for up to 16 years, for a mean of 5.64 years. Mean CCI score was low, with most participants having ≤1 comorbidity. 440 individuals developed MBI, 651 developed MCI and 230 developed dementia (see Supplemental Figure 1).

**Table 2.** Baseline characteristics of participants.

| Characteristic | N total = 4865 |
| --- | --- |
| Age at first visit, mean (SD) | 73.48 (7.00) |
| Female, n (%) | 3157 (64.9%) |
| Race, n (%) |  |
| White | 3761 (77.3%) |
| Black/African American | 1072 (22.0%) |
| Other | 32 (0.7%) |
| Years of education, mean (SD) | 16.14 (5.32) |
| Years of follow-up, mean (SD) | 5.64 (3.92) |
| Mean CCI score (SD) | 0.28 (0.70) |

Abbreviations: number (N), standard deviation (SD), Charlson Comorbidity Index (CCI)

Figure 1 displays the distribution of baseline ACB scores. These ranged from 0-11. 63.3% of participants had a score of 0. Of those taking anticholinergic medication, 21.0% had a score of 1, 6.7% had a score of 2, and 9% had a score of 3 or greater.

**Figure 1.**
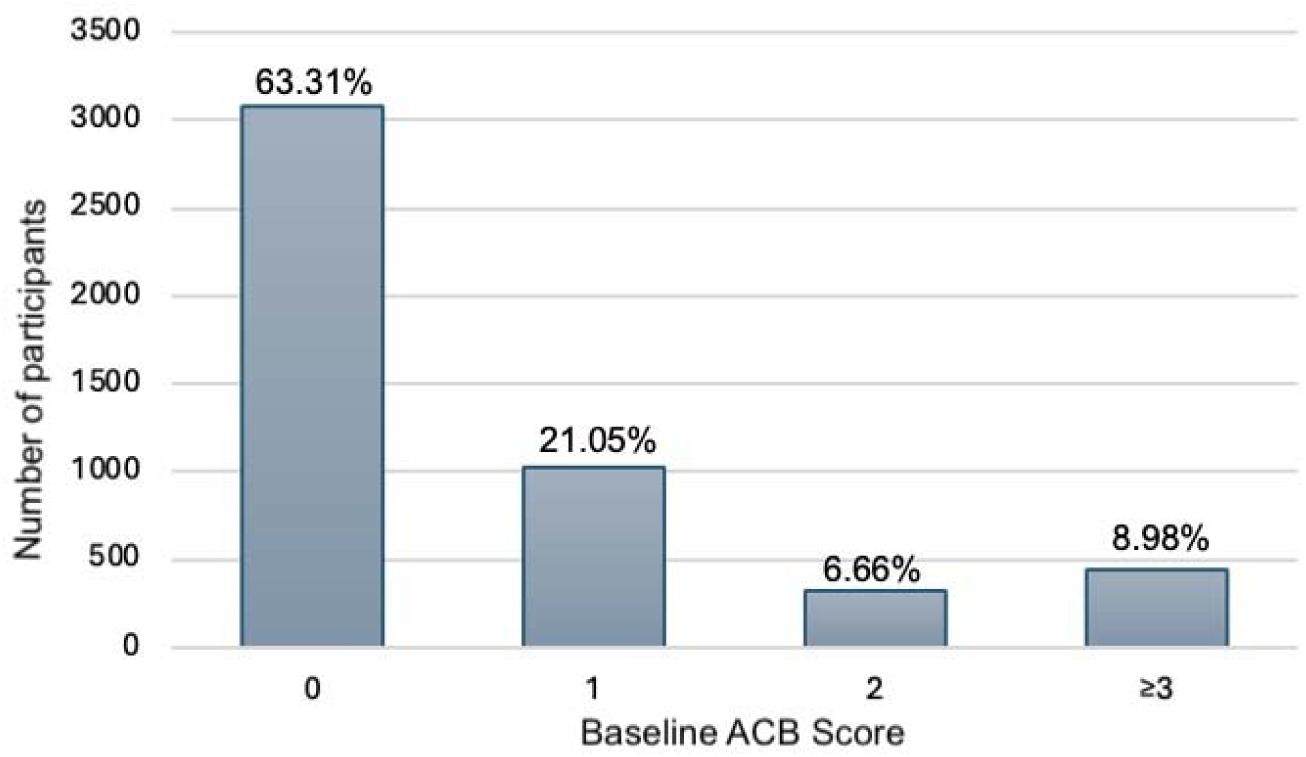
Breakdown of baseline ACB scores. The figure above each bar represents the percentage of total participants with the corresponding baseline ACB score. Abbreviations: anticholinergic burden (ACB)

To examine the impact of ACB score on the development of MBI, we plotted MBI-free survival probability in those with max ACB scores of 0, 1, 2, and 3 or greater (figure 2). Unlike in the Cox models, we used a participant’s maximum ACB score to create the Kaplan-Meier plot because it was not possible to display a survival curve with ACB as a time-varying predictor. Of 440 individuals who developed MBI, 106 (3.44%) had a maximum ACB score of 0, 105 (10.6%) had a maximum score of 1, 55 (17.0%) had a maximum score of 2, and 174 (39.8%) had a maximum score of 3 or more. A log rank test comparing the four groups was statistically significant (p=<0.001), indicating that a higher maximum total ACB score was significantly associated with a higher likelihood of developing MBI.

**Figure 2.**
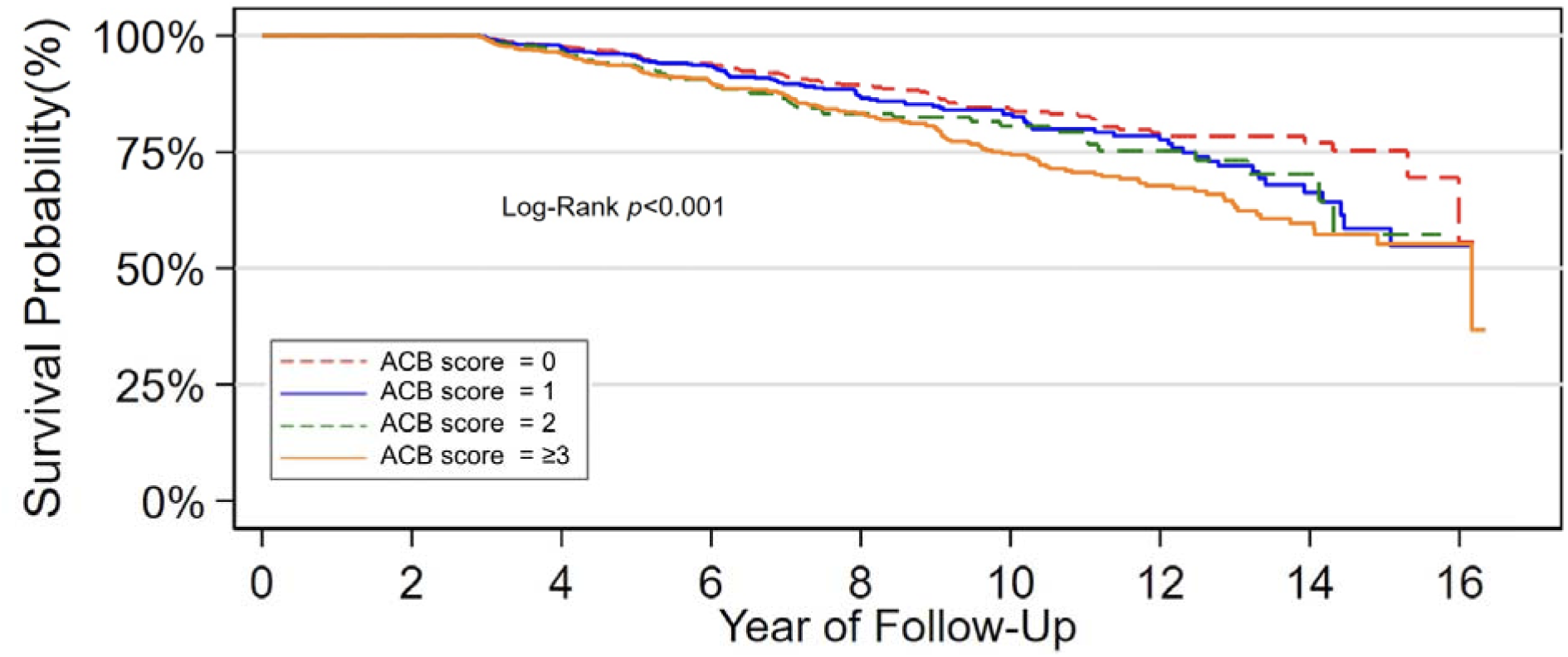
Kaplan-Meier plot of MBI-free survival. Maximum ACB score was used to facilitate the production of a Kaplan-Meier plot. A log rank test comparing the four groups was statistically significant (p=<0.001). Abbreviations: anticholinergic burden (ACB)

Table 3 shows the number of participants who reported taking selected medication and medication classes at baseline. This table includes all medications in each class that participants reported taking at baseline, not only those listed in the ACB scale. 338 (7.0%) reported taking antidepressants, those included in the ACB scale are: bupropion, fluvoxamine, trazodone, venlafaxine, amitriptyline, amoxapine, clomipramine, desipramine, doxepin, imipramine, nortriptyline, paroxetine. 15 (0.3%) reported taking antipsychotics, those included in the ACB scale are: aripiprazole, asenapine, haloperidol, iloperidone, paliperidone, loxapine, risperidone, methotrimeprazine, molindone, pimozide, chlorpromazine, clozapine, olanzapine, perphenazine, quetiapine, trifluoperazine and thioridazine. 99 (2.0%) reported taking benzodiazepines, those included in the ACB scale are: diazepam, alprazolam and clorazepate. 89 (1.8%) reported taking opiates, those included in the ACB scale are: codeine, fentanyl, meperidine, and morphine. 53 (1.1%) reported taking diphenhydramine, and 73 (1.5%) reported taking oxybutynin, both medications having an ACB score of 3. 772 (15.9%) reported taking non-aspirin NSAIDs, and 1420 (29.2%) reported taking aspirin.

**Table 3.** Number of participants who reported taking selected medication and medication classes at baseline.

| Drug class | Reported taking (%) |
| --- | --- |
| Non-aspirin NSAIDs | 772 (15.9%) |
| Opioids | 89 (1.8%) |
| Benzodiazepines | 99 (2.0%) |
| SSRIs | 169 (3.5%) |
| Non-SSRI antidepressants | 169 (3.5%) |
| Typical antipsychotics | 6 (0.1%) |
| Atypical antipsychotics | 9 (0.2%) |
| Individual drug | Reported taking (%) |
| Aspirin | 1420 (29.2%) |
| Diphenhydramine | 53 (1.1%) |
| Oxybutynin | 73 (1.5%) |

Abbreviations: Non-steroidal anti-inflammatory drugs (NSAIDs), selective serotonin reuptake inhibitor (SSRI)

Table 4 shows the hazard ratios for incident MBI. In both unadjusted and adjusted models ACB was included as a time-varying covariate. In the unadjusted model, ACB score was significantly associated with greater hazard of incident MBI (HR 1.12, 95% CI 1.05-1.19, p=<0.001). This association remained significant when adjusted for age, sex, education, and race (HR 1.11, 95% CI 1.04-1.19, p=0.001), as well as CCI score (HR 1.11, 95% CI 1.04-1.18, p=0.002). For all model parameter estimates for each covariate see supplemental table 2.

**Table 4.** Impact ACB score on the development of MBI.

| Variable | Unadjusted model |  |  | Adjusted for baseline age,<br>sex, education, race |  |  | Adjusted for baseline age,<br>sex, education, race, CCI |  |  |
| --- | --- | --- | --- | --- | --- | --- | --- | --- | --- |
|  | HR | 95% CI | p-value | HR | 95% CI | p-value | HR | 95% CI | p-value |
| ACB score | **1.12** | **1.05-1.19** | **<0.001** | **1.11** | **1.04-1.19** | **0.001** | **1.11** | **1.04-1.18** | **0.002** |

Unadjusted and unadjusted Cox proportional hazard models with co-variables are included. ACB score was treated as a time-varying predictor. Bolded figures represent statistically significant findings. Abbreviations: anticholinergic burden (ACB), hazard ratio (HR), confidence interval (CI), Charlson Comorbidity Index (CCI)

Table 5 shows the hazard ratios of taking different drugs from specific classes, or selected individual medications, as time-varying covariates regarding risk of incident MBI. Drug classes with anticholinergic activity, opioids (HR 2.12, 95% CI 1.42-3.10, p=<0.001), SSRIs (HR 2.16, 95% CI 1.45- 3.22, p=<0.001) and benzodiazepines (HR 2.19, 95% CI 1.37-3.51, p=0.001) were associated with a significantly increased hazard of incident MBI in the univariate model. Opioids, SSRIs, and benzodiazepines remained significantly associated with development of MBI in both multivariate models. Non-SSRI antidepressants and antipsychotics were not associated with incident MBI. The only individual medication significantly associated with risk of incident MBI was aspirin (HR 1.31, 95% CI 1.08-1.59, p=0.007), this significance persisted when adjusted for baseline age, sex, education, race, and CCI score.

**Table 5.** Association of drug classes and individual drugs with development of MBI. Unadjusted and unadjusted Cox proportional hazard models with co-variates are included. Medications and medication classes were treated as a time-varying predictors.

| Drug class |  |  |  |  |  |  |  |  |  |
| --- | --- | --- | --- | --- | --- | --- | --- | --- | --- |
| Variable | Unadjusted model |  |  | Adjusted for baseline age, sex, education, race |  |  | Adjusted for baseline age, sex, education, race, CCI |  |  |
|  | HR | 95% CI | p-value | HR | 95% CI | p-value | HR | 95% CI | p-value |
| Non-aspirin NSAIDs | 0.86 | 0.66-1.13 | 0.281 | 0.89 | 0.67-1.17 | 0.393 | 0.89 | 0.67-1.17 | 0.387 |
| Opioids | **2.12** | **1.42-3.19** | **<0.001** | **2.19** | **1.47-3.24** | **<0.001** | **2.06** | **1.37-3.09** | **0.001** |
| Benzodiazepines | **2.19** | **1.37-3.51** | **0.001** | **2.21** | **1.38-3.56** | **0.001** | **2.19** | **1.36-3.51** | **0.001** |
| SSRIs | **2.16** | **1.45-3.22** | **<0.001** | **2.23** | **1.50-3.34** | **<0.001** | **2.21** | **1.48-3.31** | **<0.001** |
| Non-SSRI antidepressants | 1.41 | 0.87- 2.29 | 0.165 | 1.40 | 0.86-2.29 | 0.172 | 1.395 | 0.86-2.27 | 0.180 |
| Typical antipsychotics | 2.04 | 0.29-14.57 | 0.475 | 2.197639 | 0.31-15.67 | 0.432 | 1.909315 | 0.27-13.73 | 0.520 |
| Atypical antipsychotics | $1.26 \times 10^{-14}$ | n/a | >0.999 | $9.58 \times 10^{-20}$ | n/a | n/a | $9.54 \times 10^{-20}$ | n/a | n/a |
| Individual drug |  |  |  |  |  |  |  |  |  |
| Variable | Unadjusted model |  |  | Adjusted for baseline age, sex, education, race |  |  | Adjusted for baseline age, sex, education, race, CCI |  |  |
|  | HR | 95% CI | p-value | HR | 95% CI | p-value | HR | 95% CI | p-value |
| Aspirin | **1.31** | **1.08-1.59** | **0.007** | **1.25** | **1.02-1.52** | **0.028** | **1.25** | **1.03-1.52** | **0.026** |
| Diphenhydramine | 1.04 | 0.49-2.21 | 0.908 | 1.11 | 0.53-2.35 | 0.781 | 1.11 | 0.53-2.36 | 0.776 |
| Oxybutynin | 1.55 | 0.85-2.81 | 0.154 | 1.58 | 0.87-2.88 | 0.133 | 1.57 | 0.86-2.87 | 0.138 |

Bolded figures represent statistically significant findings. Abbreviations: anticholinergic burden (ACB), hazard ratio (HR), confidence interval (CI), Charlson Comorbidity Index (CCI), non-steroidal anti-inflammatory drugs (NSAIDs), selective serotonin reuptake inhibitor (SSRI)

## 4. Conclusions

To our knowledge, this is the first study to examine the impact of anticholinergic medication exposure on the development of MBI in CU individuals. Anticholinergic exposure, as measured by the ACB scale, was significantly associated with an increased hazard of MBI in models adjusted for baseline age, sex, education, race and 10-year mortality risk. The association was dose dependent, with increasing ACB ratings associated with increasing likelihood of developing MBI.

One possible mechanism for this is the cholinergic anti-inflammatory pathway, in which the vagus nerve modulates inflammation through the parasympathetic nervous system. Acetylcholine is thought to down-regulate inflammation by reducing the production of pro-inflammatory cytokines.^38^ There have recently been significant advances in the use of muscarinic receptor agonists, coupled with a peripheral muscarinic receptor antagonist, in the management of psychotic symptoms in schizophrenia, as well as an increasing interest in studying how the anti-inflammatory effect of cholinergic agents may improve neuropsychiatric symptoms of dementia.^39,40^

When looking at anticholinergic drug classes, SSRIs were associated with an increased risk of developing MBI, while non-SSRIs were not. This conflicts with evidence that SSRIs are neuroprotective as they are associated with delayed progression from MCI to AD in individuals with depressive symptoms, and fewer amyloid plaques in mouse models and humans.^36,41^ A more recent observational study of patients with dementia reported several antidepressants including SSRIs and non-SSRIs were associated with faster cognitive decline. They addressed the issue of indication bias by performing a repeat analysis limited to only those with no history of depression.^42^ If participants were prescribed antidepressants to treat pre-existing NPS, it is possible that protopathic bias (i.e., when a medication is prescribed to treat symptoms of a disease before the disease is diagnosed, leading to an incorrect interpretation that the medication itself caused the disease in question) strengthened the association of SSRIs with risk of MBI. Given both SSRIs and non-SSRIs share clinical indications, these findings go against the argument that SSRIs may be associated with increased MBI risk as a result of protopathic bias. However, non-SSRIs also have distinct non-psychiatric indications, namely insomnia, decreased appetite, pain, and smoking cessation. As we do not know the indications for which each medication was prescribed, it is difficult to draw firm conclusions on the impact of protopathic bias on the relationship of antidepressants to MBI risk.

Opioids were associated with an increased risk of MBI. The association between opiates and dementia risk is controversial, with studies showing conflicting results.^43,44^ The impact of opioids may be exaggerated as some of the side effects of opiates can mimic NPS, namely mood and perceptual disturbances.

Though benzodiazepines are not generally thought of as anticholinergic medications, alprazolam and diazepam are both included in the ACB scale (supplemental table 1). Benzodiazepines were associated with greater hazard of MBI. This may partly explain the results of prior studies examining the impact of benzodiazepine use on dementia risk, though these studies have shown mixed results. Billioti de Gage et al. found benzodiazepines associated with an increased risk of AD, while Imfield et al. found no association.^45,46^

Study strengths include the large sample size, systematic assessment of medications and diagnoses, systematic follow-up, use of a well-validated measure of anticholinergic burden, and a mean duration of 5.64 years follow-up. ACB was examined as a time-varying covariate in the Cox models, allowing us to more accurately capture the effect of a participant’s cumulative anticholinergic burden.

Several limitations are notable. As with any longitudinal study, some participants were lost to follow-up and as a result we may have not captured all cases of MBI that developed. We did not directly assess for MBI, but mapped NPI-Q to the MBI-C using a previously described method and therefore may not be accurately capturing MBI. Not all MBI-C items are reflected in the NPI-Q, making it likely that we are under-capturing MBI. NPI-Q assesses symptoms in the past 30 days whereas NPS must be persistent over a 6-month period to meet MBI criteria. To mitigate this, we only considered MBI to be present if there was persistence of symptoms on the NPI-Q over two successive annual visits. Given this, we may both be over-capturing MBI if symptoms are not persistent for 6 months between visits, and under-capturing MBI as we are doubling the time a participant is required to experience NPS from 6 months to 12. In terms of medication data, current medication is captured but not past medication use. We also don’t capture aspects of actual medication use (e.g., regular or *as needed*), and there is no assessment of adherence. Some medications in the ACB are over the counter and, as a result, may not be as accurately reported. This may have affected our findings as our results indicate that MBI risk increases in a dose-dependent manner. The ACB scale was last updated in 2012, 8 years prior to the end of our data collection, as a result, we may not be capturing anticholinergic exposures to newer medications. By using the ACB, which encompasses medications with multiple indications, we endeavored to reduce the effect of certain medications being prescribed to treat pre-existing neuropsychiatric symptoms (i.e. antidepressants, antipsychotics), however protopathic bias cannot be completely mitigated given these medications are included in the ACB scale. That these medications had mixed effects on the likelihood of developing MBI, suggest the effect of protopathic bias may be limited. ADRC participants are not a true representation of the general population. They are overall more educated, and healthier (as evidenced by the relatively low percentage of participants taking antidepressants or benzodiazepines), and there is a lower proportion of non-White participants. As a result, participants may have a different anticholinergic burden and risk of cognitive impairment.

We report that anticholinergic exposure contributes to the development of MBI, a known precursor to dementia. Future prospective studies should be done to look at how cumulative anticholinergic exposure impacts the incidence of MBI, and should include how long prior to the development of MBI the medication was started, how long each medication was taken, at what dose. Anticholinergic medication prescription is a modifiable risk factor, and we advise that MBI risk be considered when weighing up the use of anticholinergic medication in older adults.

## Data Availability

All data produced in the present study are available upon reasonable request to the authors

## Author contributions

CEF, JML, CGL, and PBR conceptualized the project. CEF drafted the manuscript. CEF drafted the tables and figure 1. JML performed the statistical analysis and drafted figure 2. JML, CGL, PBR, and ZI edited the manuscript and approved the final version.

## Acknowledgments

The NACC database is funded by NIA/NIH Grant U24 AG072122. NACC data are contributed by the NIA-funded ADRCs: P30 AG062429 (PI James Brewer, MD, PhD), P30 AG066468 (PI Oscar Lopez, MD), P30 AG062421 (PI Bradley Hyman, MD, PhD), P30 AG066509 (PI Thomas Grabowski, MD), P30 AG066514 (PI Mary Sano, PhD), P30 AG066530 (PI Helena Chui, MD), P30 AG066507 (PI Marilyn Albert, PhD), P30 AG066444 (PI David Holtzman, MD), P30 AG066518 (PI Lisa Silbert, MD, MCR), P30 AG066512 (PI Thomas Wisniewski, MD), P30 AG066462 (PI Scott Small, MD), P30 AG072979 (PI David Wolk, MD), P30 AG072972 (PI Charles DeCarli, MD), P30 AG072976 (PI Andrew Saykin, PsyD), P30 AG072975 (PI Julie A. Schneider, MD, MS), P30 AG072978 (PI Ann McKee, MD), P30 AG072977 (PI Robert Vassar, PhD), P30 AG066519 (PI Frank LaFerla, PhD), P30 AG062677 (PI Ronald Petersen, MD, PhD), P30 AG079280 (PI Jessica Langbaum, PhD), P30 AG062422 (PI Gil Rabinovici, MD), P30 AG066511 (PI Allan Levey, MD, PhD), P30 AG072946 (PI Linda Van Eldik, PhD), P30 AG062715 (PI Sanjay Asthana, MD, FRCP), P30 AG072973 (PI Russell Swerdlow, MD), P30 AG066506 (PI Glenn Smith, PhD, ABPP), P30 AG066508 (PI Stephen Strittmatter, MD, PhD), P30 AG066515 (PI Victor Henderson, MD, MS), P30 AG072947 (PI Suzanne Craft, PhD), P30 AG072931 (PI Henry Paulson, MD, PhD), P30 AG066546 (PI Sudha Seshadri, MD), P30 AG086401 (PI Erik Roberson, MD, PhD), P30 AG086404 (PI Gary Rosenberg, MD), P20 AG068082 (PI Angela Jefferson, PhD), P30 AG072958 (PI Heather Whitson, MD), P30 AG072959 (PI James Leverenz, MD).

PBR has received research grants from the National Institutes of Aging, Alzheimer’s Clinical Trials Consortium, Richman Family Precision Medicine Center of Excellence on Alzheimer’s Disease, Eisai, Functional Neuromodulation, and Lilly. CGL has received research funding from the NIH, the Alzheimer’s Association, NFL Benefits, Functional Neuromodulation Ltd, Bright Focus Foundation and private donors. JML receives funding from NIH/NIA grants as a co-investigator.

## Conflict of interest

PBR has received honoraria from Lilly, GLG, Leerink, Acadia, Medalink, Novo Nordisk, Noble Insights, TwoLabs, Otsuka, Lundbeck, Acadia, MedaCorp, ExpertConnect, HMP Global, Sinaptica, Worldwide Clinical Trials, Medscape, and Neurology Week. CGL has served as paid consultant or advisor for Astra-Zeneca, Glaxo-Smith Kline, Eisai, Novartis, Forest, Supernus, Adlyfe, Takeda, Wyeth, Lundbeck, Merz, Lilly, Pfizer, Genentech, Elan, NFL Players Association, NFL Benefits Office, Zinfandel, BMS, Abvie, Janssen, Orion, Servier, Astellas, SVB Leerink, Roche, Avanir, Karuna, Maplight, Axsome, GIA, GW Research Limited, Merck, EXCIVA GmbH, Otsuka, IntraCellular Therapies, Medesis, BMS, Abbvie. ZI has served as advisor/consultant for the Canadian Agency for Drugs and Technology in Health, Eisai, Lilly, Lundbeck/Otsuka, Novo Nordisk, and Roche. CEF and JML have no conflicts of interest to declare.

## Notes

### Funding Statement

This study was funded by the Richman Family Precision Medicine Center of Excellence of for Alzheimers Disease, National Institute on Aging (P30AG066507)

### Author Declarations

All contributing Alzheimers Disease Research Centers are required to obtain informed consent from their participants and maintain their own local IRB reviews and ethical approvals prior to submitting data to the National Alzheimers Coordinating Center.

