## Supplemental tables and figures for "The impact of anticholinergic burden on the development of mild behavioral impairment"

**Supplemental table 1**

Medications that make up the anticholinergic burden scale

| Medication with an ACB score of 1 | Medication with an ACB score of 2 | Medication with an ACB score of 3 |
|---|---|---|
| Alimemazine | Amantadine | Amitriptyline |
| Alverine | Belladonna | Amoxapine |
| Alprazolam | Carbamazepine | Atropine |
| Aripiprazole | Cyclobenzaprine | Benztropine |
| Asenapine | Cyproheptadine | Brompheniramine |
| Atenolol | Loxapine | Carbinoxamine |
| Bupropion | Meperidine | Chlorpheniramine |
| Captopril | Methotrimeprazine | Chlorpromazine |
| Cetirizine | Molindone | Clemastine |
| Chlorthalidone | Nefopam | Clomipramine |
| Cimetidine | Oxcarbazepine | Clozapine |
| Clidinium | Pimozide | Darifenacin |
| Clorazepate | | Desipramine |
| Codeine | | Dicyclomine |
| Colchicine | | Dimenhydrinate |
| Desloratadine | | Diphenhydramine |
| Diazepam | | Doxepin |
| Digoxin | | Doxylamine |
| Dipyridamole | | Fesoterodine |
| Disopyramide | | Flavoxate |
| Fentanyl | | Hydroxyzine |
| Furosemide | | Hyoscyamine |
| Fluvoxamine | | Imipramine |
| Haloperidol | | Meclizine |
| Hydralazine | | Methocarbamol |
| Hydrocortisone | | Nortriptyline |
| Iloperidone | | Olanzapine |
| Isosorbide | | Orphenadrine |
| Levocetirizine | | Oxybutynin |
| Loperamide | | Paroxetine |
| Loratadine | | Perphenazine |
| Metoprolol | | Promethazine |
| Morphine | | Propantheline |
| Nifedipine | | Propiverine |
| Paliperidone | | Quetiapine |
| Prednisone | | Scopolamine |
| Quinidine | | Solifenacin |
| Ranitidine | | Thioridazine |
| Risperidone | | Tolterodine |
| Theophylline | | Trifluoperazine |
| Trazodone | | Trihexyphenidyl |
| Triamterene | | Trimipamine |
| Venlafaxine | | Trospium |
| Warfarin | | |

**Supplemental table 2** Model parameter estimates for each covariate used in our ACB score analysis

| **Variable** | **Adjusted for baseline age, gender, education, race** | | | **Adjusted for baseline age, gender, education, race, CCI** | | |
|---|---|---|---|---|---|---|
| | HR | 95% CI | p-value | HR | 95% CI | p-value |
| ACB score | **1.11** | **1.04-1.19** | **0.001** | **1.11** | **1.04-1.18** | **0.002** |
| Baseline age | **1.04** | **1.03-1.06** | **<0.001** | **1.04** | **1.03-1.06** | **<0.001** |
| Sex (female) | 0.87 | 0.71-1.06 | 0.169 | 0.88 | 0.72-1.08 | 0.215 |
| Education (years) | 1.01 | 0.99-1.03 | 0.185 | 1.01 | 0.99-1.03 | 0.200 |
| Race (white) | **1.51** | **1.13-2.01** | **0.006** | **1.52** | **1.14-2.03** | **0.005** |
| CCI score | | | | 1.07 | 0.99-1.16 | 0.107 |

Unadjusted and unadjusted Cox proportional hazard models with co-variates are included. ACB score was treated as a time-varying predictor. Bolded figures represent statistically significant findings. Abbreviations: anticholinergic burden (ACB), hazard ratio (HR), confidence interval (CI), Charlson Comorbidity Index (CCI).

**Supplemental Figure 1**

Diagnoses of MCI, MBI, neuropsychiatric symptoms and dementia by visit

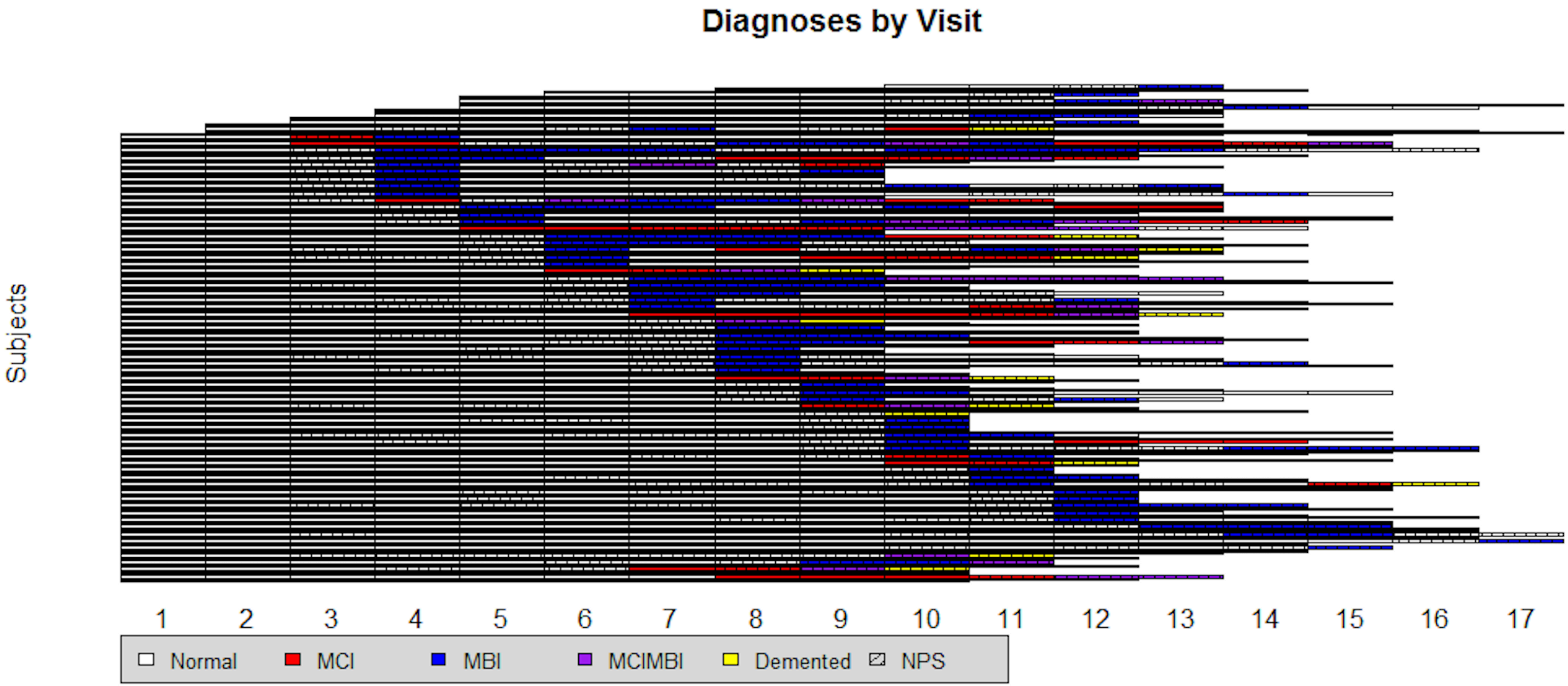


Only those subjects with 9 or more annual visits were included. Abbreviations: Mild cognitive impairment (MCI), mild behavioral impairment (MBI), neuropsychiatric symptoms (NPS)
